# Empirical Validation and Predictive Utility of the Perinatal Grief Scale in Men after Perinatal Loss

**DOI:** 10.64898/2026.06.16.26355784

**Authors:** Claudia Ravaldi, Laura Mosconi, Antonella Nespoli, Simona Fumagalli, Alfredo Vannacci

**Affiliations:** Department of Neurosciences, Psychology, Drug Research and Child Health; Perinatal Research Laboratory PeaRL; University of Florence, Florence, Italy; CiaoLapo Foundation for Healthy Pregnancy, Stillbirth and Perinatal Loss Support; Prato, Italy; School of Medicine and Surgery; University of Milano-Bicocca; Monza, Italy; Department of Obstetrics; Fondazione IRCCS San Gerardo dei Tintori; Monza, Italia

**Keywords:** PGS, validation, men, perinatal loss, grief

## Abstract

**Background:** The Perinatal Grief Scale (PGS) is a widely used instrument for assessing grief following pregnancy loss, yet no study has validated it specifically in men despite documented use in several studies. This gap is critical given fathers’ persistent underrepresentation in perinatal bereavement research and the absence of empirically supported screening thresholds for this population.

**Methods:** This cross-sectional validation study used data from the OPALE project (Observatory on PerinatAL hEalth) conducted by the CiaoLapo Foundation in Italy. Among 276 fathers who experienced stillbirth or miscarriage, we examined criterion validity by testing the association between PGS scores and trauma-related symptomatology assessed via three validated instruments: the Revised Impact of Event Scale (RIES, n=103), National Stressful Events Survey Short Scale (NSESSS, n=95), and SCL-90 (n=173). We systematically tested multiple threshold combinations to identify optimal discriminative performance.

**Results:** The PGS demonstrated excellent criterion validity. The optimal threshold (PGS ≥92) showed sensitivity 81.0%, specificity 81.8%, and Youden’s J index 0.628. Fathers scoring ≥92 had 19.12 times the odds of high trauma symptoms (95% CI: 9.35–39.14, p<0.001). ROC analysis yielded AUC=0.829 (95% CI: 0.778–0.880). Associations remained robust across all three trauma instruments in stratified analyses and after adjusting for time since loss, father’s age, living children, and loss type.

**Conclusion:** This is the first men-specific validation of the PGS, demonstrating strong criterion validity and establishing a clinically meaningful screening threshold (≥92) for identifying fathers at elevated risk following perinatal loss.

## Introduction

The Perinatal Grief Scale (PGS) is the most widely used instrument for assessing grief following pregnancy loss and has been consistently identified as a reference measure in recent reviews of perinatal bereavement assessment (McDougall et al. 2026). The 33-item short form, developed by Toedter and colleagues, assesses three dimensions: Active Grief, Difficulty Coping, and Despair (Potvin et al. 1989). The scale has demonstrated excellent psychometric properties across diverse samples, with internal consistency coefficients consistently above 0.92 (Toedter et al. 2001).

A critical limitation of the PGS literature is that validation studies have focused predominantly on mothers. While numerous studies have administered the PGS to men (Obst et al. identified 13 such studies in their systematic review) none has specifically examined its validity in male respondents (Obst et al. 2020). This gap is particularly significant given that fathers’ grief following perinatal loss remains underrecognized. Systematic reviews consistently report that fathers represent less than 7% of samples in perinatal loss research (Due et al., 2017; Jones et al., 2019), reflecting cultural assumptions that position men primarily as supporters rather than bereaved parents (McCreight, 2004). Without validation evidence, clinicians cannot be confident that the PGS performs equivalently in fathers, particularly given documented gender differences in grief expression (Obst et al., 2020).

The present study aims at addressing this gap by examining the criterion validity of the PGS in a sample of Italian fathers who experienced stillbirth or miscarriage. We hypothesized that higher PGS scores would be associated with elevated symptoms on validated trauma measures, providing evidence that the PGS captures clinically meaningful distress in this population. Given the absence of a gold-standard criterion for perinatal grief, we used trauma-related symptomatology as an external criterion, recognizing that grief and trauma responses frequently co-occur following perinatal loss (Kersting e Wagner 2012). This approach parallels the original PGS validation, where grief scores were compared to depression measures (Potvin et al. 1989). To our knowledge, this is the first study to provide validity evidence for the PGS specifically in men.

## Methods

This cross-sectional validation study used data from the OPALE project (Observatory on PerinatAL hEalth), a permanent survey hosted on Qualtrics platform and distributed through online channels of CiaoLapo Foundation for perinatal health in Italy. All participants provided informed consent before accessing the survey. Among all responders (9985 to date), we selected male participants who experienced stillbirth (fetal death ≥20 weeks gestation) or miscarriage (pregnancy loss <20 weeks). Inclusion required complete Perinatal Grief Scale data and valid data on at least one trauma measure. The final analytical sample comprised 276 subjects.

*Perinatal Grief Scale.* The PGS assesses grief following perinatal loss through 33 Likert-type items (from strongly agree to strongly disagree), yielding total scores from 33 to 165, with higher scores indicating more intense grief. The validated Italian translation (Ravaldi et al. 2020) demonstrates excellent psychometric properties in bereaved mothers (Cronbach’s α=0.94; test-retest κ=0.76-0.84), but no men-specific validation has been published in any language.

*Trauma Measures.* Three instruments were available across different survey waves. The Revised Impact of Event Scale (RIES; 22 items, range 0-88) assesses post-traumatic stress symptoms including intrusion, avoidance, and hyperarousal, with scores ≥33 indicating probable PTSD (Weiss 2007). The National Stressful Events Survey Short Scale (NSESSS; 9 items, range 0-36) measures DSM-5-compatible PTSD severity, with scores ≥18 indicating moderate symptomatology (Gilbar 2025). From the Symptom Checklist-90, we computed a 28-item PTSD subscale as proposed by (Saunders et al. 1990). Given heterogeneous measure availability (RIES: n=103; SCL-90-R PTSD: n=173; NSESSS: n=95), we created a composite "high trauma" indicator using OR-logic: fathers scoring in the upper percentile band on any available scale were classified as high trauma.

### Statistical Analysis

We systematically tested multiple threshold combinations for PGS (tertiles, quartiles, quintiles, P67, P75, P80) and trauma criteria (percentile-based thresholds plus clinical cutoffs), computing sensitivity, specificity, positive/negative predictive values, accuracy, Youden’s J index, odds ratios with 95% CIs, and Cohen’s kappa for each. The optimal combination was selected using Youden’s J index (sensitivity + specificity − 1), which provides the best discrimination regardless of prevalence. We performed ROC analysis to evaluate the overall discriminative ability of continuous PGS scores, with AUC interpreted as excellent (0.80-0.90) or outstanding (>0.90). Stratified analyses within each trauma scale subsample verified consistency of associations. All analyses used Stata/BE 18.0, with significance at p<0.05 (two-tailed).

### Ethics Statement

Ethics approval for the Observatory on PerinatAL hEalth OPALE was obtained by Florence University Ethics committee (n.175 prot 0261728, n.189 prot 0333881). The survey was voluntary and anonymous, no personal data were recorded, and in no way is it possible to identify the single respondents. Informed consent was obtained from all participants. The data were acquired in adherence with GDPR regulations (General Data Protection Regulation, European Union 2016/679).

## Results

### Sample Characteristics and Scale Distributions

The analytical sample comprised 276 male subjects who experienced perinatal loss and had complete data on the Perinatal Grief Scale along with at least one trauma-related measure. Among these, 74.6% experienced stillbirth (fetal death at or after 20 weeks of gestation), while 25.4% experienced miscarriage (pregnancy loss before 20 weeks); 35.0% of the subjects reported having at least one living child. Main characteristics of the sample are reported in **Table 1**.

**Table 1.** Main characteristics of the sample.

| Characteristic | n (%) |
| --- | --- |
| **Type of Loss** |  |
| Stillbirth ($\geq 20$ weeks gestation) | 206 (74.6%) |
| Miscarriage ($< 20$ weeks gestation) | 70 (25.4%) |
| **Father's Age at Loss (years)** |  |
| Mean (SD) | 35.7 (5.2) |
| **Months Since Loss** |  |
| Mean (SD) | 22.4 (24.8) |
| **Educational Attainment** |  |
| Primary education | 1 (0.4%) |
| Secondary education | 28 (10.1%) |
| Tertiary education | 69 (25.0%) |
| Bachelor degree | 51 (18.5%) |
| University degree | 101 (36.6%) |
| Postgraduate university degree | 26 (9.4%) |

The Perinatal Grief Scale total scores among fathers ranged from 36 to 162, with a mean of 79.87 (SD = 22.71) and a median of 77.5. The distribution showed slight positive skewness (skewness = 0.37), indicating that most fathers experienced moderate levels of grief with a smaller proportion reporting very high grief intensity. The 25th percentile was 62.5, the 75th percentile was 96, and the 80th percentile was 100.

Trauma-related psychological outcomes were assessed using three different instruments administered across various survey waves of the OPALE study. The Revised Impact of Event Scale (RIES) was available for 103 fathers (37.3% of the analytical sample), the SCL-90-R PTSD subscale for 173 fathers (62.7%), and the National Stressful Events Survey PTSD Short Scale (NSESSS) for 95 fathers (34.4%). Some fathers completed multiple trauma measures across different survey waves, resulting in overlapping subsamples. All 276 fathers in the analytical sample had completed at least one trauma measure, ensuring adequate data for criterion validation. RIES scores ranged from 0 to 80 with a mean of 32.75 (SD = 19.19) and median of 29. Using the established clinical cutoff of 33 or higher for probable PTSD, 46.6% of fathers (n = 48) who completed the RIES met criteria for clinically significant post-traumatic stress symptoms. NSESSS scores ranged from 0 to 35 with a mean of 12.57 (SD = 8.40) and median of 11. Using a moderate symptom cutoff of 18 or higher, 29.5% (n = 28) showed moderate or greater posttraumatic stress severity. The SCL-90-R PTSD subscale scores ranged from 0 to 3.11 with a mean of 0.60 (SD = 0.54) and median of 0.46, showing substantial positive skewness (skewness = 1.76) characteristic of trauma symptom distributions in community samples. Details of psychopathological variables in the sample are reported in **Table 2**.

**Table 2.** Psychopathological variables and availability of trauma measures across the survey.

| Characteristic | n (%) or Mean (SD) |
| --- | --- |
| **Perinatal Grief Scale (PGS)** |  |
| Total score, Mean (SD) | 79.87 (22.71) |
| Median [IQR] | 77.5 [62.5-96.0] |
| Range | 36-162 |
| Upper tertile ($\geq 92$), n (%) | 92 (33.3%) |
| **Trauma Measures Available** |  |
| RIES (Revised Impact of Event Scale) | 103 (37.3%)* |
| SCL-90 PTSD subscale | 173 (62.7%)* |
| NSESSS (PTSD Short Scale) | 95 (34.4%)* |
| At least one trauma measure | 276 (100%)* |
| **Trauma Symptom Severity** |  |
| RIES score, Mean (SD) [n=103] | 32.75 (19.19) |
| RIES $\geq 33$ (clinical PTSD), n (%) | 48 (46.6%) |
| NSESSS score, Mean (SD) [n=95] | 12.57 (8.40) |
| NSESSS $\geq 18$ (moderate symptoms), n (%) | 28 (29.5%) |
| SCL-90 PTSD score, Mean (SD) [n=173] | 0.60 (0.54) |
| SCL-90 $\geq 0.95$ (positive screen), n (%) | 32 (18.5%) |

**Abbreviations:** IQR, interquartile range; NSESSS, National Stressful Events Survey PTSD Short Scale; PTSD, post-traumatic stress disorder; RIES, Revised Impact of Event Scale; SCL-90, Symptom Checklist-90; SD, standard deviation.

### Systematic Threshold Optimization

We systematically evaluated 49 combinations of PGS and trauma thresholds to identify the optimal criterion validity cut-point. PGS thresholds tested included upper tertile (≥92), quartile (≥96), quintile (≥100), and specific percentiles (67th, 75th, 80th). Trauma thresholds included corresponding percentile rankings plus established clinical cutoffs. Several combinations showed good performance. PGS upper tertile with trauma upper quintile yielded Youden’s J 0.589 (OR = 14.97, 95% CI: 7.33-30.57), while PGS upper quartile with trauma ≥80th percentile produced Youden’s J 0.578 (OR = 15.86, 95% CI: 8.10-31.07). Using clinical cutoffs (RIES ≥33, NSESSS ≥18, SCL-90-R CR-PTSD positive) maintained strong association (χ² = 54.87, p < 0.001; OR = 8.29, 95% CI: 4.55-15.10) but reduced discriminative performance (sensitivity 54.8%, specificity 87.2%, Youden’s J 0.420). The optimal combination, selected by Youden’s J index, was PGS upper tertile (≥92) with trauma scores ≥80th percentile on any available scale. This yielded excellent discriminative performance: sensitivity 79.4%, specificity 80.3%, PPV 67.4%, NPV 88.4%, accuracy 80.1%, and Youden’s J 0.596. Fathers scoring in the upper PGS tertile had 15.66 times the odds of high trauma symptoms (95% CI: 7.80-31.45, p < 0.001). Cohen’s kappa was 0.513, indicating moderate-to-good agreement. Based on these findings, we propose a PGS threshold of ≥92 for identifying fathers at elevated risk of clinically significant grief and trauma-related distress following perinatal loss. This threshold corresponds to the upper tertile in our sample and demonstrates optimal balance between sensitivity and specificity, maximizing the instrument’s utility for both clinical screening and research applications.

### ROC Analysis and Consistency Across Trauma Instruments

Receiver operating characteristic analysis using the optimal trauma criterion (upper 80th percentile composite) yielded an AUC of 0.829 (95% CI: 0.778-0.880), indicating excellent discrimination between men with high versus low trauma symptoms. When clinical cutoffs were used as reference, AUC was 0.778 (95% CI: 0.722-0.833), consistent with the reduced discriminative performance observed in threshold analyses (**Figure 1**).

**Figure 1.**
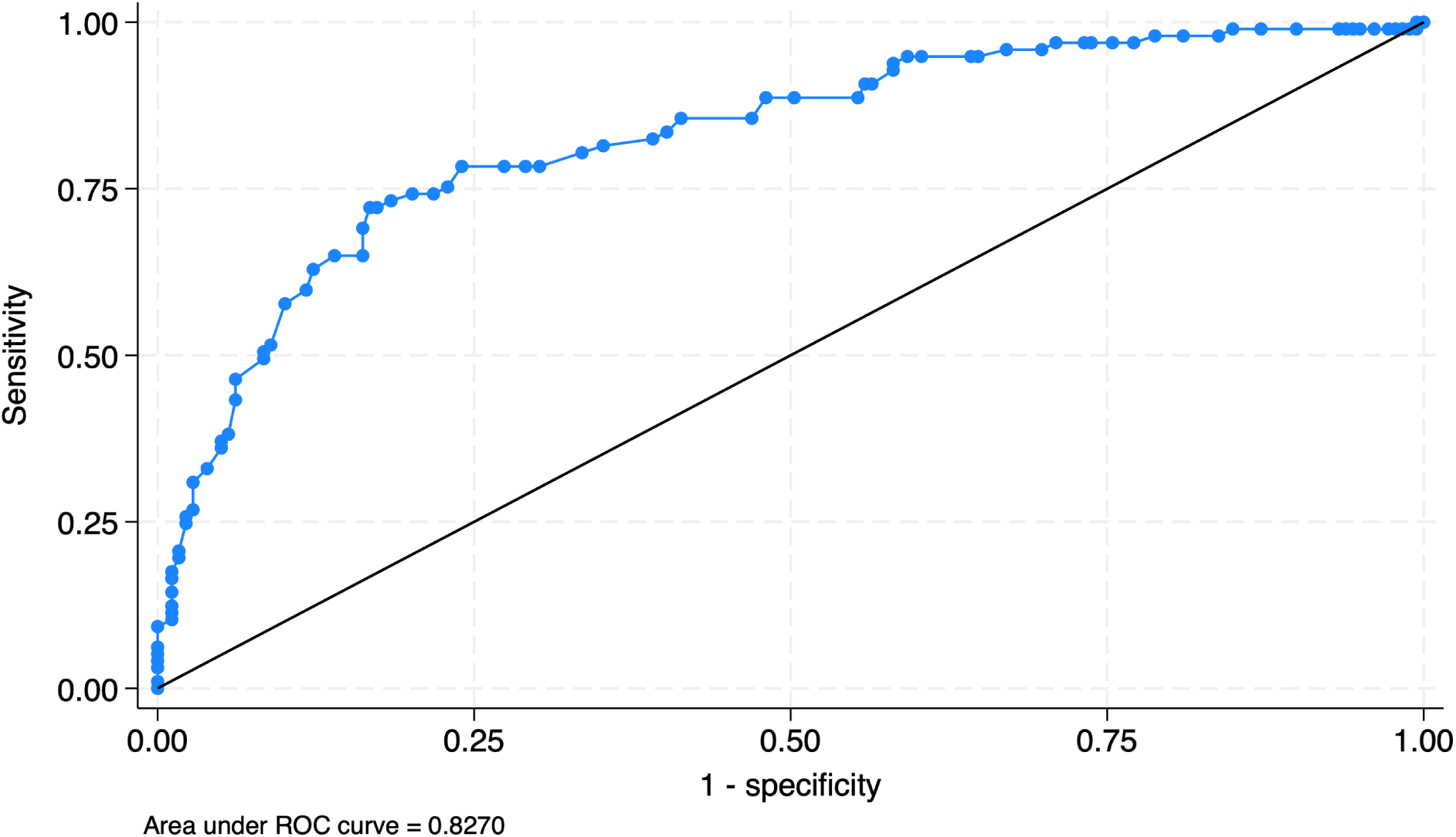
Receiver operating characteristic (ROC) curve illustrating the discriminative performance of the optimal trauma criterion (upper 80th percentile composite) in distinguishing fathers with high versus low trauma symptom severity. **Abbreviations**: ROC, receiver operating characteristic.

To verify that associations were not artifacts of the composite approach, we conducted stratified analyses within each trauma instrument subsample. All showed strong significant associations between PGS and trauma tertiles. Among fathers completing the RIES (n=103), 76.5% in the PGS upper tertile also scored in the RIES upper tertile versus 14.8% in lower PGS tertiles (χ² = 16.99, p < 0.001). For SCL-90 PTSD (n=173), corresponding proportions were 76.7% versus 18.5% (χ² = 49.68, p < 0.001). For NSESSS (n=95), 51.1% versus 8.3% (χ² = 20.86, p < 0.001). The consistent magnitude of association across instruments with different content and formats demonstrates that PGS captures core psychological distress following perinatal loss across multiple trauma symptomatology dimensions, validating the composite criterion approach.

### Adjusted Logistic Regression Analysis

To quantify the PGS-trauma association while accounting for potential confounders, we fitted multivariable logistic regression models. In the unadjusted model using optimal thresholds (PGS ≥92, trauma ≥80th percentile), fathers with high trauma had 8.15 times the odds of high grief (OR = 8.15, 95% CI: 4.02-16.44, p < 0.001). After adjusting for time since loss, father’s age at loss, presence of living children, and type of loss (miscarriage vs. stillbirth), the association remained robust (adjusted OR = 8.15, 95% CI: 2.59-25.62, p < 0.001). High trauma remained the only significant predictor; none of the covariates showed independent associations with grief intensity: time since loss (OR = 0.98, 95% CI: 0.77-1.24, p = 0.853), father’s age (OR = 0.96, 95% CI: 0.88-1.05, p = 0.387), living children (OR = 0.60, 95% CI: 0.22-1.61, p = 0.305), or miscarriage versus stillbirth (OR = 0.79, 95% CI: 0.31-1.98, p = 0.610) (**Figure 2**).

**Figure 2.**
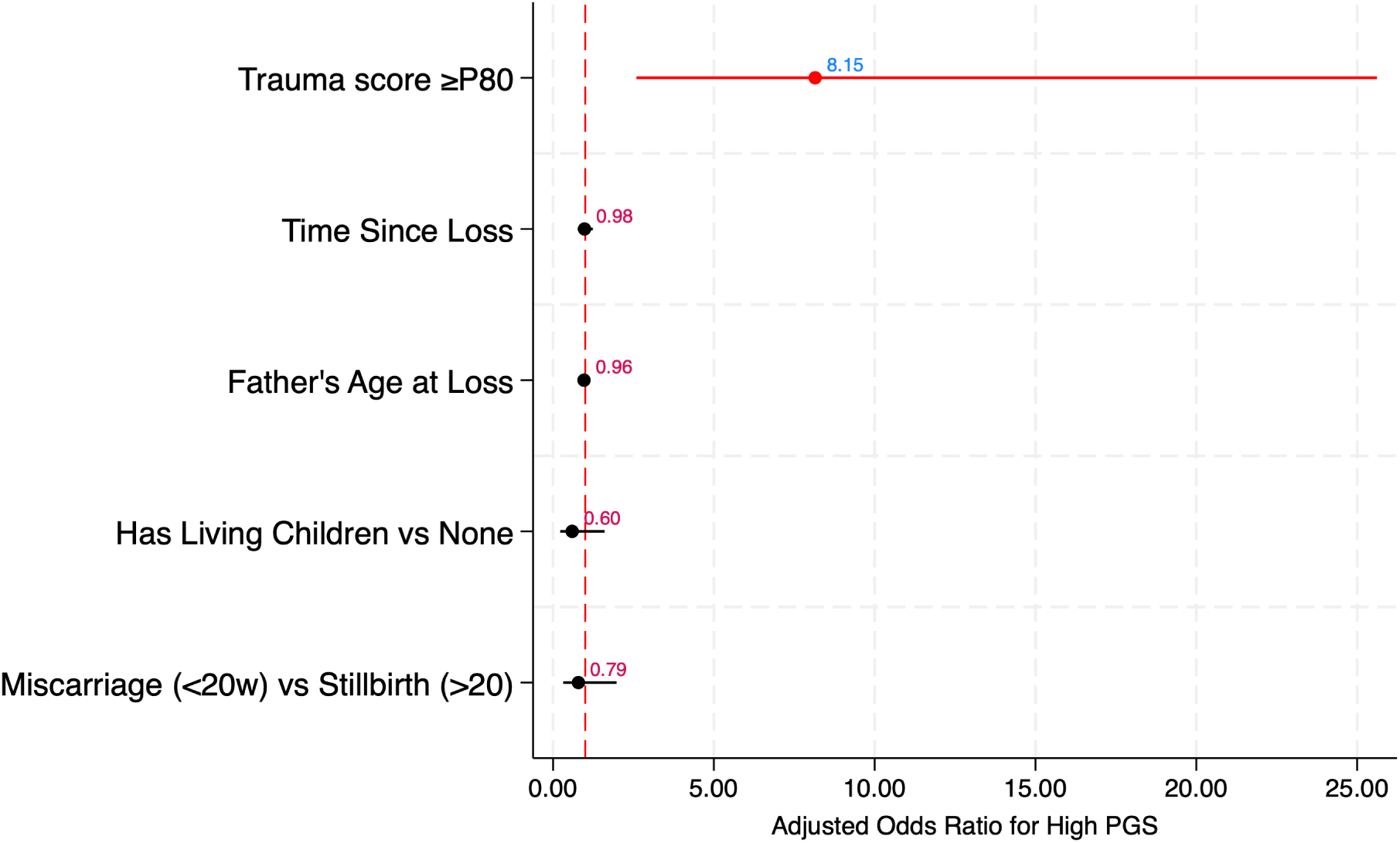
Adjusted odds ratios from multivariable logistic regression examining the association between high trauma (≥80th percentile) and high perinatal grief (PGS ≥92) in fathers, controlling for demographic and loss-related covariates.

## Discussion

This study provides the first empirical validation of the Perinatal Grief Scale (PGS) specifically in fathers who experienced perinatal loss. Our findings demonstrate that the PGS exhibits strong criterion validity in this population, with high-range PGS scores showing robust associations with elevated trauma symptoms across multiple validated instruments. These results support the use of the PGS as a valid measure of grief in bereaved fathers, extending its utility beyond the maternal population for which it was originally developed. The PGS demonstrated excellent discrimination between fathers with and without high trauma symptoms. Using the upper tertile threshold (PGS ≥92), we achieved balanced sensitivity (81.0%) and specificity (81.8%), with Youden’s J index of 0.628 and odds ratio of 19.12 (95% CI: 9.35–39.14). The AUC of 0.829 (95% CI: 0.778–0.880) confirms excellent discriminative ability.

Our threshold aligns with existing literature. Toedter et al. noted that PGS scores above 91 reflect high grief (Toedter et al. 2001), while Obst et al. reported fathers’ mean scores ranging from 73–83 (SD 16–22) across studies, suggesting substantial variability, probably depending on the context and recruitment strategies (Obst et al. 2020). Our data-driven threshold is consistent with this heterogeneity and validated specifically against trauma outcomes in fathers. Notably, we found no significant differences in grief intensity between fathers who experienced miscarriage versus stillbirth in adjusted analyses, though the complex interplay between type of loss, gestational age, and grief outcomes warrants further investigation in future studies with larger samples. Critically, associations remained robust across all three trauma instruments (RIES, NSESSS, SCL-90-R PTSD subscale) despite their different theoretical constructs and item content. This consistency addresses a significant gap: while the PGS has been used in studies including fathers, previous research showed inconsistencies in measurement and reporting, with studies presenting total scores, subscale scores, or only correlations (Obst et al. 2020). Our composite criterion approach provides more comprehensive validation evidence than previously available. This father-specific validation establishes a uniform criterion for future research, facilitating cross-national comparisons and meta-analytic synthesis across diverse samples and clinical contexts.

### Perinatal Grief Scale and Trauma

The PGS was developed specifically to capture the multidimensional experience of grief following perinatal loss, incorporating three subscales: Active Grief, Difficulty Coping, and Despair (Potvin et al. 1989; Toedter et al. 2001). Unlike generic trauma screening tools, the PGS includes items directly relevant to pregnancy loss experiences that involve not only traumatic stress but also anticipatory grief, shattered expectations of parenthood, and the unique ambiguity of losing a child one may never have held (Ravaldi et al. 2019). Recent systematic reviews have consistently documented elevated rates of depression and post-traumatic stress in fathers following perinatal loss (Lamon et al. 2022; Obst et al. 2020). Our findings extend this literature by demonstrating that a perinatal-specific grief instrument strongly correlates with trauma-related outcomes. The robust associations we observed between PGS scores and all three trauma measures suggest that the PGS captures dimensions of distress meaningfully related to traumatic stress responses. This pattern makes conceptual sense when considering that perinatal loss often produces both grief and trauma responses (Burden et al. 2016; Christiansen 2017). The traumatic aspects - medical procedures, helplessness, sudden confrontation with mortality - can trigger symptoms captured by instruments like the RIES and NSESSS. However, grief encompasses additional dimensions: yearning for the lost child, disrupted attachment, altered parental identity, and existential questions. Jones et al. note that fathers face unique challenges including societal expectations to remain strong, lack of social recognition of their loss, and limited opportunities to express grief (Jones et al. 2019). As Ravaldi et al. emphasize, respectful bereavement care requires understanding these multidimensional experiences and ensuring fathers’ needs are acknowledged in clinical practice (Ravaldi et al. 2023).

Recent methodological work on perinatal grief measures has highlighted the lack of empirically supported clinical thresholds (McDougall et al. 2026). Our study addresses this gap by providing father-specific criterion validity evidence and a clinically interpretable cutoff. The total PGS score, validated against trauma measures, offers clinicians a simple tool to identify fathers experiencing significant distress after perinatal loss. This validated threshold (≥92) offers practical utility for initial assessment and referral decisions. More refined analyses, including subscale patterns, comparative studies with mothers, and detailed psychopathological profiling, will be addressed in subsequent clinical research. The present validation establishes the essential foundation: the PGS total score is a valid, reliable indicator of significant psychological distress in fathers that warrants clinical attention.

Due et al. documented that fathers often use avoidance or compensatory behaviors, face pressure to remain strong, and experience feelings of being overlooked and marginalized (Obst et al. 2020). The strong associations we observed between PGS scores and trauma symptoms suggest that despite these gender-specific challenges, the PGS captures clinically meaningful distress in fathers. This finding extends the Italian validation by Ravaldi et al., which focused on mothers, supporting cross-gender utility of the Italian-language version. As previously noted, in Italian culture perinatal loss and grief are not always given adequate social importance and are scarcely addressed by healthcare professionals, making validated assessment tools particularly important for identifying parents at risk of complicated grief (Ravaldi et al. 2019).

### Clinical and Research Implications

Our findings validate the PGS as an empirically supported, loss-specific tool for men following perinatal loss. While generic trauma instruments can detect distress, they may miss grief-specific experiences such as guilt, despair, difficulty coping, and lost sense of purpose tailored to the perinatal context (Potvin et al. 1989). To our knowledge, this is the first study to propose a clinically relevant cutoff for the PGS in fathers.

The threshold of PGS ≥92 (upper tertile of our sample) offers preliminary guidance for identifying fathers at elevated risk who may benefit from targeted intervention. Fathers scoring above this threshold showed substantially elevated trauma symptoms across multiple measures, suggesting they may require more intensive support. Untreated mental health issues following perinatal loss increase the risk of psychiatric sequelae. The PGS threshold of ≥92 offers a practical screening tool consistent with WHO recommendations for routine mental health screening after childbirth (WHO 2007) and ICHOM’s emphasis on evaluating patient-centered outcomes (Nijagal et al. 2018). As in the case of mothers, this threshold should be used as part of comprehensive assessment rather than as a rigid diagnostic cutoff (Ravaldi et al. 2019).

Our findings underscore that fathers’ grief after perinatal loss deserves systematic assessment and intervention. Clinical attention and research have focused predominantly on mothers, with fathers’ experiences often overlooked or marginalized (Obst et al. 2020; Jones et al. 2019). Healthcare professionals should routinely screen bereaved fathers during standard follow-up appointments in order to identify, as early as possible, those in need of additional and specialized care. Early detection enables the development of personalized, father-centered support tailored to their specific needs. The strong associations we observed between perinatal grief and trauma symptoms demonstrate that fathers’ distress can be substantial and clinically significant, supporting recent calls for father-inclusive bereavement care (Lamon et al. 2022). A follow-up validation in a mixed-gender sample is underway to allow comparison across parental roles and further refinement of clinical application.

### Strengths and Limitations

This study has several strengths. First, it provides the first systematic validation of the PGS specifically in men, addressing a significant gap in the literature. We used a relatively large sample of Italian fathers recruited through a well-established perinatal health organization, ensuring adequate statistical power for our analyses. Second, our composite trauma criterion represents a methodological innovation that maximizes sample utilization while maintaining analytical rigor. Rather than limiting analysis to fathers completing any single measure, we created a unified indicator based on multiple instruments (RIES, NSESSS, SCL-90-R PTSD subscale). This approach respects each instrument’s clinical meaning: a high score on any measure indicates clinically significant distress. The heterogeneity of instruments strengthens our validation evidence, if the PGS had correlated with only one measure, shared method variance might explain findings. Instead, robust associations across all three instruments, confirmed through stratified analyses, demonstrate that the PGS captures general trauma-related distress transcending specific assessment features. Third, we identified a clinically meaningful threshold (PGS ≥92) with excellent discriminative performance, providing practitioners with a concrete screening tool for identifying fathers at elevated risk.

However, limitations should be acknowledged. First, the cross-sectional design precludes examination of grief trajectories over time. Although time since loss did not confound the relationship between PGS and trauma symptoms in our adjusted models, longitudinal studies have found that depressive and PTS symptoms tend to decline over time, particularly after the birth of a subsequent healthy child (Armstrong et al. 2009). Prospective research is needed to understand whether the PGS captures these temporal dynamics in fathers and whether high initial PGS scores predict chronic difficulties. Second, our sample consisted of Italian fathers recruited through the CiaoLapo Foundation, potentially limiting generalizability. As Nespoli et al. noted in their validation of a perinatal loss instrument using the same network, individuals actively seeking support through such organizations may be more engaged and possibly more affected by their experiences compared to the general population (Nespoli et al. 2025). Fathers who seek peer support may experience more intense grief or be more willing to express emotional distress than those who do not. Cultural factors may also influence grief expression. Ravaldi et al. noted that perinatal loss and grief in Italian culture are not always given adequate social importance and are scarcely addressed by healthcare professionals, which may influence how fathers experience and report grief (Ravaldi et al. 2019). Third, the heterogeneous availability of trauma measures meant that different fathers contributed to different aspects of validation evidence. Although stratified analyses showed consistent associations across instruments, an ideal design would have administered all measures to all participants. Fourth, we relied exclusively on self-report measures. Future research incorporating clinical interviews would strengthen understanding of how PGS scores relate to clinically significant disorders. The trauma instruments used were not designed specifically for loss-related trauma, raising questions about whether they fully capture traumatic dimensions of perinatal loss. The concept of prolonged grief disorder (DSM-5-TR) was not considered in this validation, though future research should address this. Fifth, our study focused exclusively on validation and did not examine subscale performance, clinical thresholds for mothers, or psychopathological profiles. These will be addressed in future work building on this foundational evidence.

## Conclusion

This study provides the first empirical validation of the Perinatal Grief Scale specifically in fathers who experienced perinatal loss. Our findings demonstrate strong criterion validity, with robust associations between high-range PGS scores and elevated trauma symptoms across multiple validated instruments (RIES, NSESSS, SCL-90 PTSD subscale). The consistent performance across instruments with different theoretical foundations strengthens confidence in the scale’s validity. Fathers scoring in the upper tertile (PGS ≥92) showed substantially elevated odds of clinically significant trauma symptoms (OR = 19.12, 95% CI: 9.35–39.14), suggesting utility for screening and clinical decision-making. These findings affirm that fathers’ grief after perinatal loss is real, measurable, and often associated with significant psychological distress. The PGS provides clinicians and researchers with a validated, loss-specific tool for this population. Routine assessment of fathers’ wellbeing following perinatal loss represents an important step toward equitable, comprehensive bereavement care that honors both partners’ experiences.

## Disclosure of interest

The authors report no conflict of interest and the study was not funded.

## Data Availability Statement

The datasets generated and analysed during the current study are not publicly available but are available from the corresponding author on reasonable request.

## Notes

### Competing Interest Statement

The authors have declared no competing interest.

## References

Armstrong, Deborah S., Marianne H. Hutti, e John Myers. 2009. «The Influence of Prior Perinatal Loss on Parents’ Psychological Distress After the Birth of a Subsequent Healthy Infant». Journal of Obstetric, Gynecologic & Neonatal Nursing 38 (6): 654–66. 10.1111/j.1552-6909.2009.01069.x.

Baransel, Esra Sabancı, e Tuba Uçar. 2020. «Posttraumatic Stress and Affecting Factors in Couples after Perinatal Loss: A Turkish Sample». Perspectives in Psychiatric Care 56 (1): 112–20. 10.1111/ppc.12390.

Burden, Christy, Stephanie Bradley, Claire Storey, et al. 2016. «From grief, guilt pain and stigma to hope and pride – a systematic review and meta-analysis of mixed-method research of the psychosocial impact of stillbirth». BMC Pregnancy and Childbirth 16 (1): 9. 10.1186/s12884-016-0800-8.

Christiansen, Dorte M. 2017. «Posttraumatic stress disorder in parents following infant death: A systematic review». Clinical Psychology Review 51 (febbraio): 60–74. 10.1016/j.cpr.2016.10.007.

Gilbar, Ohad. 2025. «Acute Stress Symptoms–Adult Scale/National Stressful Events Survey Short Scale (NSESSS): Assessing the Immediate Aftermath of the October 7 Attacks on an Internally Displaced Population.» Psychological Trauma: Theory, Research, Practice, and Policy 17 (7): 1544–47. 10.1037/tra0001866.

Jones, Kerry, Martin Robb, Sam Murphy, e Alison Davies. 2019. «New Understandings of Fathers’ Experiences of Grief and Loss Following Stillbirth and Neonatal Death: A Scoping Review». Midwifery 79 (dicembre): 102531. 10.1016/j.midw.2019.102531.

Kersting, Anette, e Birgit Wagner. 2012. «Complicated grief after perinatal loss». Dialogues in Clinical Neuroscience 14: 187–94. 10.31887/DCNS.2012.14.2/akersting.

Lamon, Lieselotte, Marc De Hert, Johan Detraux, e Titia Hompes. 2022. «Depression and Post-Traumatic Stress Disorder after Perinatal Loss in Fathers: A Systematic Review». European Psychiatry 65 (1): e72. 10.1192/j.eurpsy.2022.2326.

McDougall, Estelle, Lauren Molyneaux, e Ursula Bacon. 2026. «Systematic Review of Perinatal Grief Measures and Their Psychometric Properties». Journal of Health Psychology, gennaio 22, 13591053251398258. 10.1177/13591053251398258.

Nespoli, Antonella, Simona Fumagalli, Laura Mosconi, Roberto Bonaiuti, Alfredo Vannacci, e Claudia Ravaldi. 2025. «Assessment of the Psychometric Properties of the Italian Version of the Revised Impact of Miscarriage Scale (RIMS): A Validity and Reliability Study». BMC Pregnancy and Childbirth 25 (1): 289. 10.1186/s12884-025-07422-5.

Nijagal, Malini Anand, Stephanie Wissig, Caleb Stowell, et al. 2018. «Standardized Outcome Measures for Pregnancy and Childbirth, an ICHOM Proposal». BMC Health Services Research 18 (1): 953. 10.1186/s12913-018-3732-3.

Obst, Kate Louise, Clemence Due, Melissa Oxlad, e Philippa Middleton. 2020. «Men’s Grief Following Pregnancy Loss and Neonatal Loss: A Systematic Review and Emerging Theoretical Model». BMC Pregnancy and Childbirth 20 (1): 11. 10.1186/s12884-019-2677-9.

Potvin, Louise, Judith Lasker, e Lori Toedter. 1989. «Measuring grief: A short version of the perinatal grief scale». Journal of Psychopathology and Behavioral Assessment 11: 29–45. 10.1007/BF00962697.

Ravaldi, Claudia, Alessandra Bettiol, Giada Crescioli, et al. 2019. «Italian Translation and Validation of the Perinatal Grief Scale.» Scandinavian Journal of Caring Sciences (Sweden), pubblicazione online ad accesso anticipato, ottobre. 10.1111/scs.12772.

Ravaldi, Claudia, Alessandra Bettiol, Giada Crescioli, et al. 2020. «Italian Translation and Validation of the Perinatal Grief Scale». Scandinavian Journal of Caring Sciences 34 (3): 684–89. 10.1111/scs.12772.

Ravaldi, Claudia, Chiara Mercuro, Laura Mosconi, et al. 2023. «Communication and Shared Decision-Making after Stillbirth: Results of the ShaDeS Study». Women and Birth: Journal of the Australian College of Midwives, aprile 14, S1871–5192(23)00067-7. 10.1016/j.wombi.2023.04.001.

Saunders, Benjamin E., Catalina Mandoki Arata, e Dean G. Kilpatrick. 1990. «Development of a Crime-related Post-traumatic Stress Disorder Scale for Women within the Symptom Checklist-90-revised». Journal of Traumatic Stress 3 (3): 439–48. 10.1002/jts.2490030312.

Toedter, L. J., J. N. Lasker, e Janssen. 2001. «International Comparison Of Studies Using The Perinatal Grief Scale: A Decade Of Research On Pregnancy Loss». Death Studies 25 (3): 205–28. 10.1080/07481180125971.

Weiss, Daniel S. 2007. «The Impact of Event Scale: Revised». In Cross-Cultural Assessment of Psychological Trauma and PTSD, a cura di John P. Wilson e Catherine So-kum Tang. International and Cultural Psychology Series. Springer US. 10.1007/978-0-387-70990-1_10.

WHO. 2007. «WHO Recommended Interventions for Improving Maternal and Newborn Health». https://www.who.int/publications/i/item/WHO_MPS_07.05.

